# Single-cell RNA sequencing reveals a novel cell type and immunotherapeutic targets in papillary thyroid cancer

**DOI:** 10.1101/2021.02.24.21251881

**Authors:** Zhengshi Wang, Youlutuziayi Rixiati, Wenli Jiang, Chen Ye, Caiguo Huang, Chuangang Tang, Zhiqiang Yin, Binghua Jiao

## Abstract

Papillary thyroid cancer (PTC) is the most common thyroid malignancy. Although PTC usually has a favorable prognosis, some aggressive PTC subtypes and lymph node (LN) metastasis contribute to high rates of recurrence and poor clinical outcomes. We analyzed single-cell RNA sequencing (scRNA-seq) data from 15 samples, including primary tumors of PTC, metastatic LNs, and paracancerous tissues. After quality filtering, 28,205 cells were detected. Of these, 13,390 cells originated from 7 tumor tissues, 2,869 cells from 2 metastatic LNs, and 11,945 cells from 6 paracancerous tissues. The increase in the proportion of CD4^+^ Tregs may be a key factor responsible for the immunosuppressive property of PTC. A novel cell type was identified, named Protective EGR1^+^CD4^+^ T cell, which might be antagonistic to the CD4^+^ Tregs and inhibit the formation of the immunosuppressive microenvironment and tumor immune evasion. Inhibitory checkpoints TIGIT and CD96 were found to be better targets than PD-1 for immune therapy in PTC patients with LN metastasis. For PTC patients without LN metastasis, however, PD-1, TIGIT, and CD96 could be suitable targets of immunotherapy. These findings would contribute to the further understanding of molecular mechanisms resulting in occurrence and development of PTC, and provide a theoretical rationale for targeted therapy and immunotherapy.

## Introduction

According to the GLOBOCAN database, there were an estimated 586,202 new cases of thyroid cancer worldwide in 2020 ^1^. The global incidence was up to 13.1 per 100,000 people, ranking 11th in occurrence rate. The most common pathological type of thyroid cancer is papillary thyroid cancer (PTC), accounting for approximately 85% of all cases ^2^. Although PTC usually has a favorable prognosis, some aggressive PTC subtypes (e.g. tall cell variant and Hürthle cell cancer) and lymph node (LN) metastasis contribute to high rates of recurrence and poor clinical outcomes ^3, 4^. For PTC patients with a high risk of recurrence, radioiodine treatment could be used to improve their prognosis ^5^. However, some PTC patients would develop into radioiodine-refractory PTC, which in turn lead to low survival rate ^6, 7^. Therefore, a deeper understanding of molecular and cellular mechanisms of PTC would help in the development of therapeutic strategies.

Currently, efforts to study the tumor progression and metastatic process of thyroid cancer have mainly focused on the analysis of cancer cells using genetic aberrations^8, 9, 10, 11^. However, the tumor progression and metastasis are a complicated biological process, which were not only affected by the characteristic features of cancer cells themselves but also by the tumor microenvironment (TME) ^12, 13, 14, 15^. TME refers to a tumor pathology-related environment, comprising stromal cells, extracellular matrix (ECM), and cytokines. A comprehensive analysis of TME in PTC can reveal the key elements involved in the susceptibility of tumor-induced immunological changes, which could be employed to develop new immunotherapy strategies.

Genomic and transcriptomic studies have revealed a huge number of driver mutations, abnormal regulatory programs, and disease subtypes in major human tumors^16, 17, 18, 19^. However, the conventional bulk sequencing frequently adopted in these studies only revealed the overall biological characteristics of each tumor and lacked the ability to capture signatures in intratumoral and intercellular heterogeneity. As opposed to bulk sequencing, the emergence of single-cell RNA sequencing (scRNA-seq) provided a new opportunity to enables characterization of cell populations at the single-cell level, and made it more independent of any previous assumptions about surface markers ^20^. scRNA-seq has been applied to investigate cellular properties in various solid tumor types ^21^. However, the single-cell atlas of PTC remains to be fully revealed. Therefore, we analyzed primary PTC tumors, paracancerous tissues, and metastatic LNs using the scRNA-seq-based profiling method to better understand the intratumoral heterogeneity and complexity during the development of PTC.

## Results

### Single cell atlas and heterogeneity of PTC

A total of 15 samples from seven PTC patients were involved in this study. After quality filtering, 28,205 cells were detected. Of these, 13,390 cells originated from 7 tumor tissues, 2,869 cells from 2 metastatic LNs, and 11,945 cells from 6 paracancerous tissues (Fig. 1a and 1b). Subsequently, we partitioned the cells into 26 clusters, which were further classified into 10 major cell types based on known markers described in previous studies: B cells (CD19^+^, MS4A1^+^, CD38^+^, CD79A^+^, CD79B^+^); CD4^+^ T cells (CD3D^+^, CD4^+^); CD8^+^ T cells (CD3D^+^, CD8A^+^); endothelial cells (CD31^+^, CD34^+)^; epithelial cells (EPCAM^+^, KRT18^+^); fibroblasts (COL1A1^+^); myeloid cells (CD14^+^, CD86^+^, ITGAX^+^, CD80^+^, CD83^+^, ITGAM^+^); naive T cells (CD3D^+^, CCR7^+^); natural killer T (NKT) cells (CD3D^+^, NKG7^+^); and plasma cells (CD79A^+^, SDC1^+^) (Fig. 1c, Supplementary Fig. 1 and 2).

**Fig.1.**
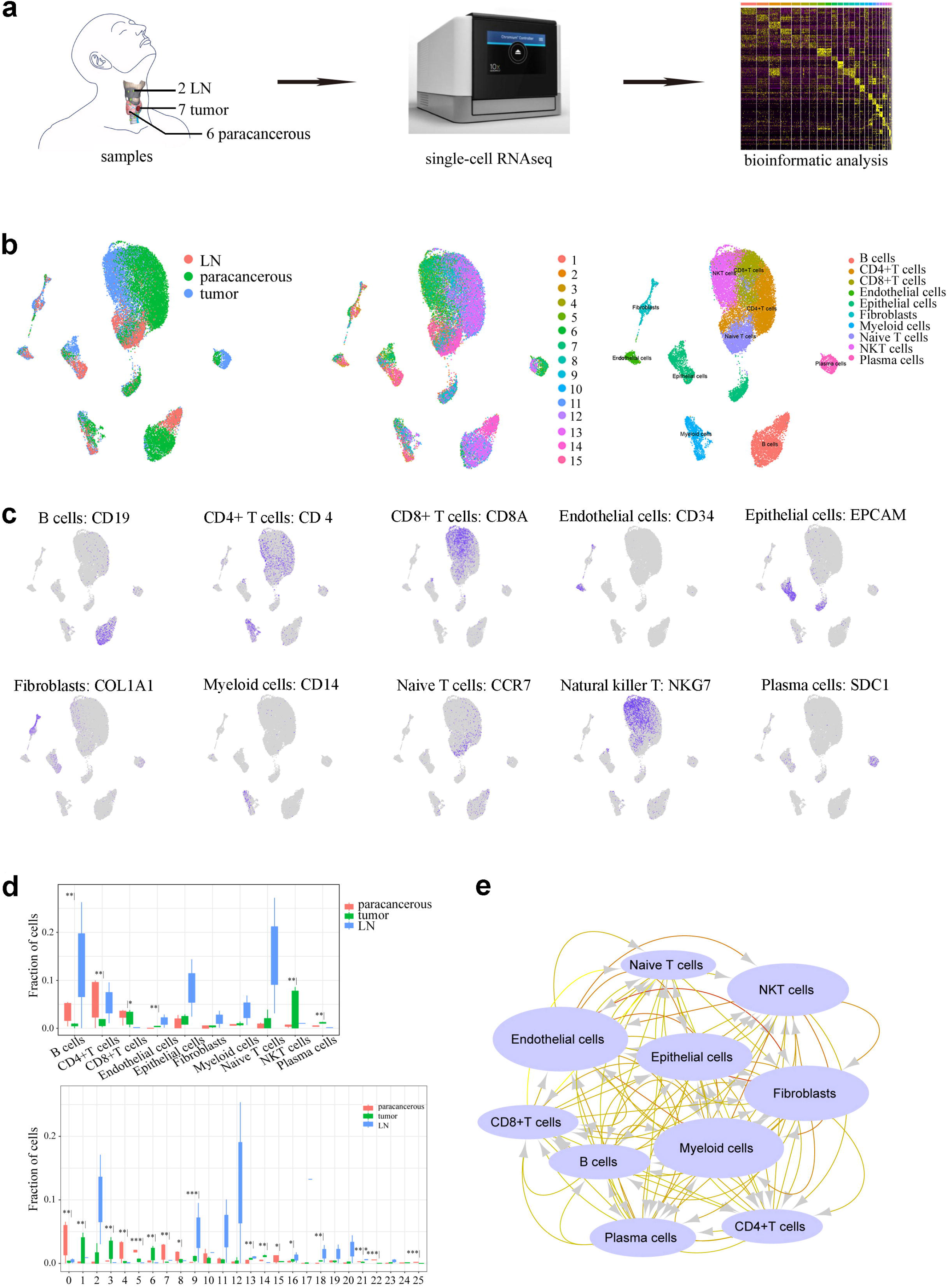
Overall design and single cell atlas in papillary thyroid carcinoma. **a** Workflow diagram showing the processing of samples. **b** UMAP plots of total cells, colored by the sample origin (tumor, metastatic LN, and paracancerous tissues). **c** Expression of cell marker genes. **d** Changes in frequency of multiple cell types and clusters in tumors and metastatic LNs. Asterisks on the left of the vertical line denote statistically significant differences between tumors and paracancerous tissues while asterisks on the right are used to show statistically significant differences between metastatic LNs and tumors. *p < 0.05, **p < 0.01, ***p < 0.001, two-tailed t-tests. **e** Interaction network constructed by CellPhoneDB. Size of circles and color of arrows represent interaction counts, and brighter color and larger size mean more interaction with other cell types.

**Fig.2.**
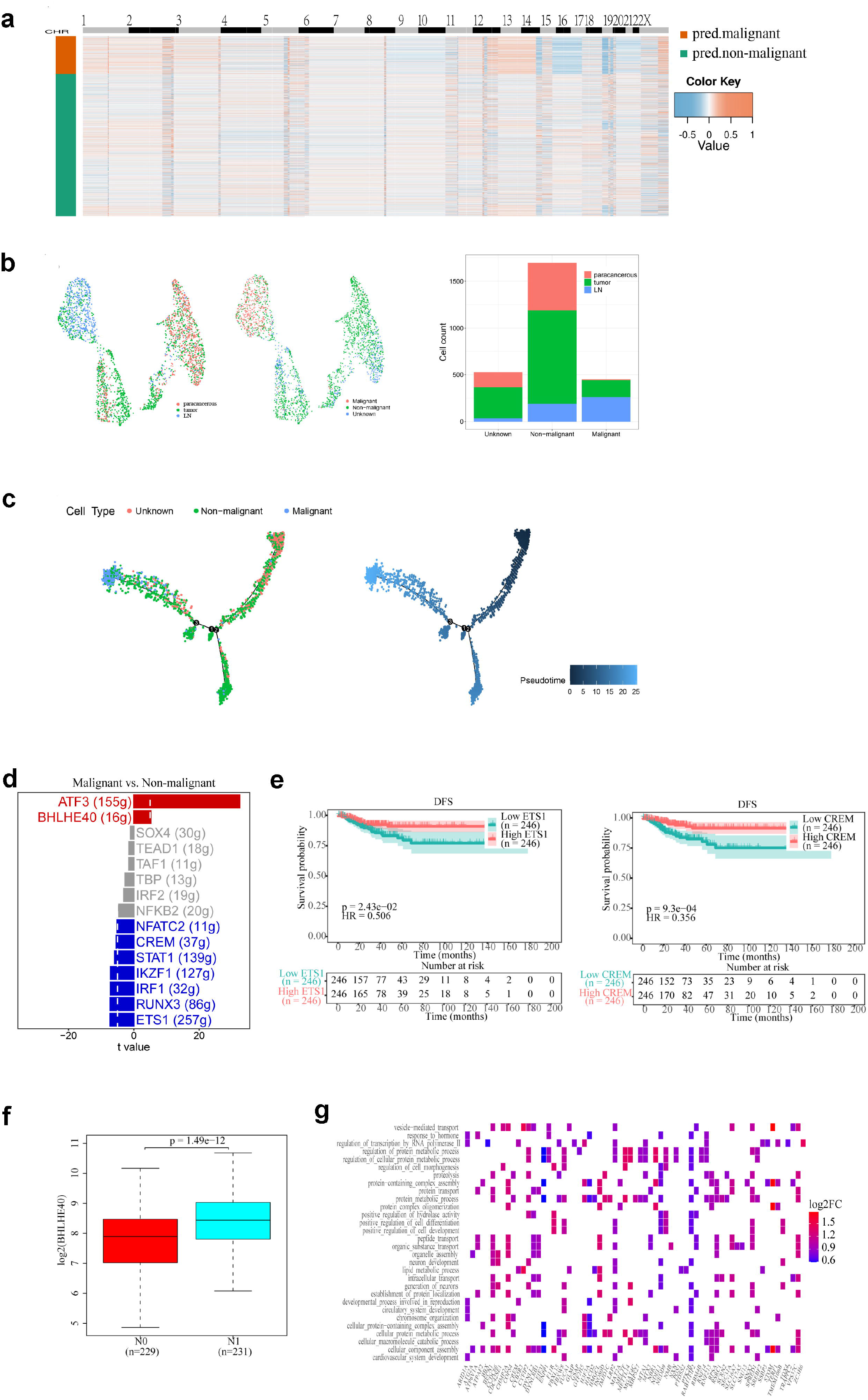
Identification of malignant cells in epithelial cells. **a** Clonal substructure of epithelial cells delineated by clustering single-cell copy number profiles inferred from scRNA-seq data by CopyKAT. **b** UMAP plots of total epithelial cells, colored by the sample origin (left and middle). Cell counts of malignant and non-malignant cells, colored by the sample origin (right). **c** Differentiation trajectory of epithelial cells, with each color coded for pseudotime (right) and clusters (left). **d** Heatmap of the area under the curve (AUC) scores of transcription factor (TF) motifs estimated by SCENIC. Shown are differentially activated motifs in malignant and non-malignant cells, respectively. **e** High level of CREM and ETS1 predicted good prognosis in the TCGA THCA cohort. Log-rank p value < 0.05 was considered as statistically significant. **f** BHLHE40 was highly expressed in LN samples (p < 0.001). **g** GO enrichment analysis of up-regulated genes in malignant lymphatic metastasis cells.

Based on the frequencies of cell types, we detected cellular landscapes in primary tumors, paracancerous tissues, and metastatic LNs, respectively. significant differences were observed in several cell types (Fig. 1d). The immune cells accounted for the major differences between primary tumors and paracancerous tissues, with the frequency of NKT cells and plasma cells increasing, and CD4^+^ T cells and B cells decreasing in the primary tumors (P<0.05). The most striking changes between metastatic LNs and primary tumors were observed in CD8^+^ T cells. At the cluster level, there were also significant differences within the same cell types. As exemplified by myeloid cells, we observed an increase in the frequency of cluster 21 and a decrease in the proportion of cluster 25 in metastatic LNs compared with primary tumors and paracancerous tissues. These results demonstrated that the disease progression might also be somehow associated with evolutionary immune cells, reflecting the existence of heterogeneity among samples and cell populations.

To conduct the interaction network of tumor microenvironment in PTC, CellphoneDB was used to calculate potential ligand-receptor pairs in cells. And Cytoscape was performed to visualize the cell interaction. We found that myeloid cells and T cell-related cells possessed more interaction pairs with other cells than others (Fig. 1e), showing the dominant roles of myeloid cells and T cells.

### Identification of malignant cells in epithelial cells

To distinguish malignant from non-malignant cells within the epithelial cells, CopyKAT was performed to identify PTC genome alterations. Despite the heterogeneity, almost all malignant cells possessed deletions from chromosomes 16 and 19 and amplifications in chromosomes 13 (Fig. 2a and 2b). We distinguished 1,679 non-malignant cells and 450 cells were identified as malignant cells. Very few malignant cells were also found in paracancerous tissues (Fig. 2b). In order to support the identification results of the malignant cells, pseudotime trajectory analysis was performed. The results showed that malignant cells were present at the end of the differentiation trajectory (Fig. 2c).

To screen for key regulons in malignant cells, we conducted SCENIC analysis and identified motifs ATF3 and BHLHE40 that were highly activated in malignant cells (Fig. 2d). In a previous study, ATF3 was found to enhance breast cancer metastasis ^36^. At the same time, decreased activity was found in CREM and ETS1. In TCGA THCA cohort, a high level of CREM and ETS1 was significantly related to a good prognosis, and BHLHE40 is highly expressed in patients with LN metastasis compared with patients without LN metastasis (Fig. 2e and 2f). These results provide potential targets for suppressing cells to possess malignant characteristics.

We further characterized the functions of differential genes between metastatic LNs and primary tumors by comparing pathway activities. Pathways involved in cellular component assembly, protein-containing complex assembly, and regulation of microtubule motor activity were relatively upregulated in LN-derived malignant cells (Fig. 2g). The results showed that malignant cells in metastatic LNs have stronger cell proliferative and invasive abilities.

### Identification of a novel T cell type

Reclustering of T cells identified 10 subclusters: CD4^+^ Tregs, Cytotoxic CD8^+^ T cells, Exhausted CD4^+^ T cells, Follicular helper (Tfh) T cels, Naïve CD4^+^ T cells, Naïve T cells; NKT cells, Pre-exhausted CD8^+^ T cells, Proliferating CD8^+^ T cells, and Protective EGR1^+^CD4^+^ T cells (Fig. 3a and 3b). In addition to the markers used above, we also found some other makers that can be used for cell population identification. For example, TYMS can be used for Pre-exhausted CD8^+^ T cells identification, and RTKN2 can be used for Pre-exhausted CD4^+^ Tregs identification (Fig. 3c, Supplementary Fig. 3). Notably, a novel cell type was identified, which was totally different from preconceived cellular definitions. The novel cell type specifically highly expressed EGR1, and we named it as Protective EGR1^+^CD4^+^T cells. The mRNA expression level and protein level of EGR1 were significantly decreased in tumor tissues compared with normal tissues (Fig. 3d). Survival analysis showed that patients with high EGR1 levels had a better prognosis than those with low EGR1 levels (Fig. 3e). Through cell frequency analysis, we found an interesting phenomenon that the proportion of CD4^+^ Tregs tended to increase with PTC disease progression and Protective EGR1^+^CD4^+^ T cells exhibited a quite opposite trend (Fig. 3f). The results were validated with TCGA data calculated using Cibersortx (Fig. 3g) and further validated by the changes of marker gene expression levels in the TCGA THCA cohort (Supplementary Fig. 4a). These findings indicated that both of CD4^+^ Tregs and Protective EGR1^+^CD4^+^ T cells may have an important impact on disease progression, and our newly identified cell type (Protective EGR1^+^CD4^+^T cells) may be antagonistic to the CD4^+^ Tregs.

**Fig.3.**
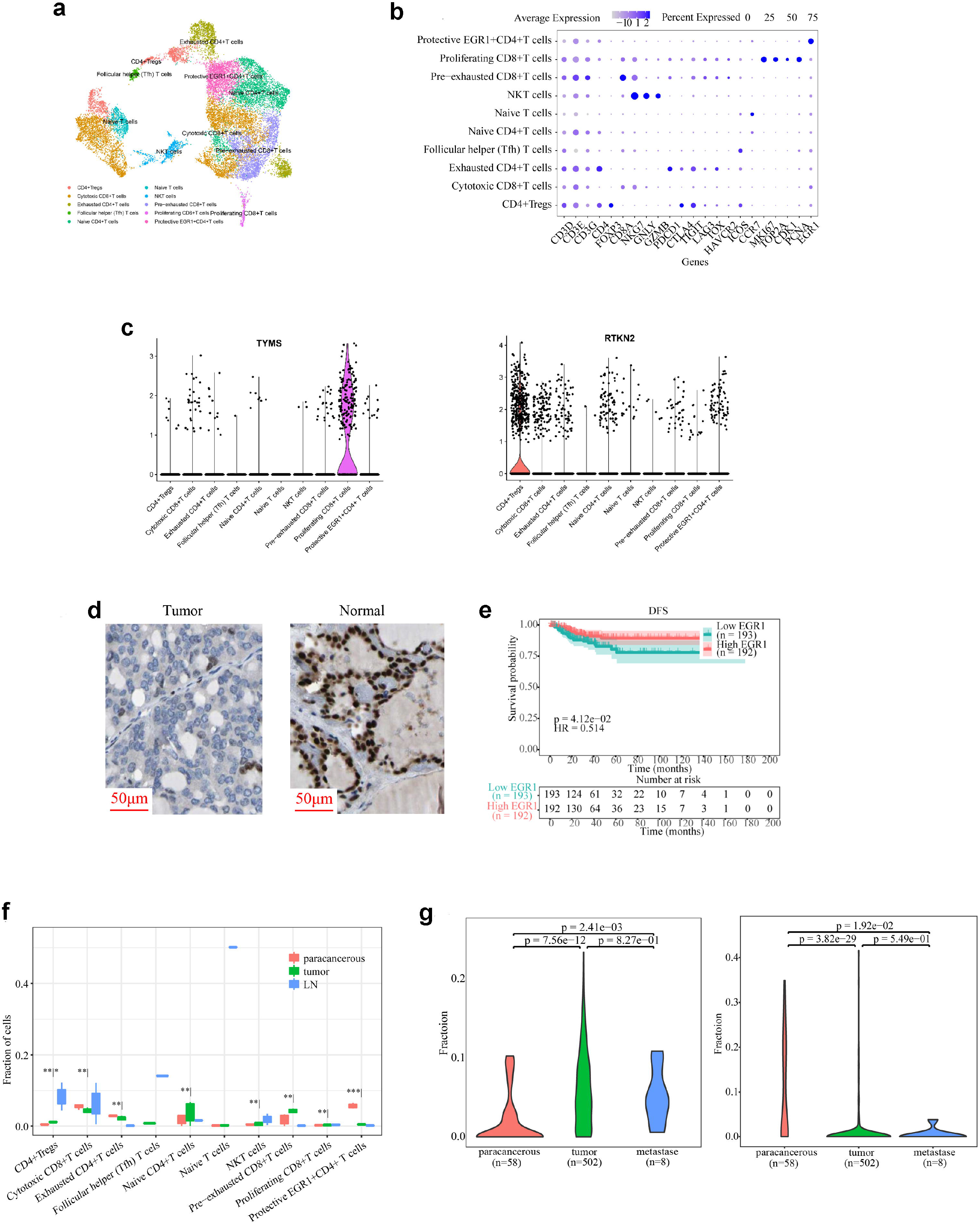
Heterogeneity of T cell populations. **a** UMAP plot of T cells color-coded by their associated cell types. **b** Dotplot of cell markers; sizes of dots represent abundance while color represents expression level. **c** Potential new cell markers. **d** The protein level of EGR1 was decreased in tumor tissues compared with normal tissues. **e** High level of EGR1 predicted good prognosis in the TCGA THCA cohort. **f** Changes in frequency of multiple cell types in T cell populations. Asterisks on the left of the vertical line denote statistically significant differences between tumors and paracancerous tissues while asterisks on the right are used to show statistically significant differences between LNs and tumors. *p < 0.05, **p < 0.01, ***p < 0.001, two-tailed t-tests. **g** Violin of cell abundance predicted from the TCGA THCA cohort by CIBERSORTx.

**Fig.4.**
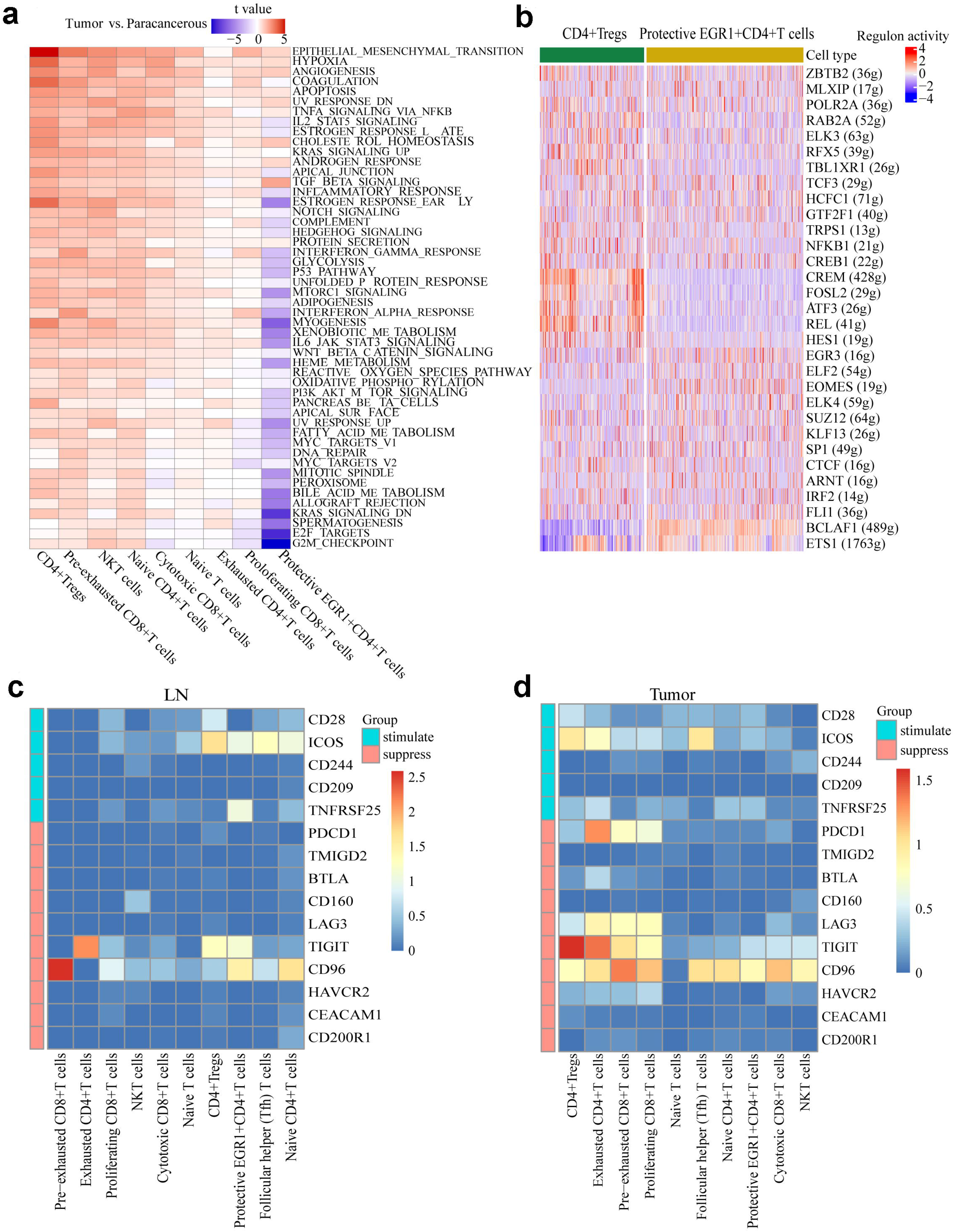
Novel cell type, named Protective EGR1^+^CD4^+^ T cells, and potential therapeutic targets. **a** Differences in 50 hallmark pathway activities scored with GSVA software. Shown are t values calculated by a linear model. **b** Heatmap of the AUC scores of TF motifs estimated by SCENIC. Shown are differentially activated motifs in CD4^+^ Tregs and Protective EGR1^+^CD4^+^ T cells, respectively. **c-d** Heatmap of positive and negative immune checkpoint expression on T cells in LNs (**c**) and tumors (**d**), respectively.

We further characterized the functions of CD4^+^ Tregs and Protective EGR1^+^CD4^+^ T cells by comparing pathway activities. In the primary tumors, CD4^+^ Tregs and Protective EGR1^+^CD4^+^ T cells exhibited vastly different signaling pathways. CD4^+^ Tregs were related to epithelial-mesenchymal transition (EMT) and hypoxia, whereas G2M checkpoint and KRAS signaling pathway were significantly down-regulated in Protective EGR1^+^CD4^+^ T cells (Fig. 4a). In the metastatic LNs, mitotic spindle and oxidative phosphorylation were also down-regulated in Protective EGR1^+^CD4^+^ T cells (Supplementary Fig. 4b). To screen for key genes related to tumorigenesis and tumor development in CD4^+^ Tregs and Protective EGR1^+^CD4^+^ T cells, we conducted SCENIC analysis and identified essential motifs in CD4^+^ Tregs and Protective EGR1^+^CD4^+^ T cells. FOSL2, ATF3, REL, and HES1 were CD4^+^ Tregs-specific motifs. BCLAF1 and ETS1 motifs were highly activated in Protective EGR1^+^CD4^+^ T cells (Fig. 4b). These results provided potential targets for inhibiting or reversing the formation of the immunosuppressive microenvironment.

The DEGs derived from Protective EGR1^+^CD4^+^ T cells were identified, including 31 up-regulated mRNAs and 50 down-regulated mRNAs in primary tumors compared with paracancerous tissues (Supplementary Fig. 4c). The DEGs derived from CD4^+^ Tregs were also identified (Supplementary Fig. 4d). Subsequently, we examined the immune checkpoints in the cell clusters (Fig. 4c and 4d). Notably, we found that TIGIT, an inhibitory checkpoint, was upregulated in Exhausted CD4^+^ T cells and CD96 was upregulated in Pre-exhausted CD8^+^ T cells in metastatic LNs. In tumor tissues, PDCD1 (PD-1) and TIGHT were significantly upregulated in Exhausted CD4^+^ T cells, and CD96 was significantly upregulated in Pre-exhausted CD8^+^ T cells. Since TIGIT, CD96, and PDCD1 (PD-1) are markers of T cell exhaustion, these data indicated that Exhausted CD4^+^ T cells and Pre-exhausted CD8^+^ T cells were exhausted in the tumor microenvironment, which was consistent with our previous cell population classification results.

### Neutrophils cells are extremely reduced in tumor

For exploring the heterogeneity among myeloid cells, 1607 myeloid cells were clustered into 5 cell subtypes based on known markers described in previous studies: macrophages, dendritic cells, monocytes, neutrophils, and myeloid-derived suppressor cells (Fig. 5a and 5b). Through cell frequency analysis, a marked decrease of neutrophils was observed in tumors while the proportion of macrophages increased continuously as the disease progressed (Fig. 5c). The trend was consistent with TCGA data calculated by cibersortx (Fig. 5d). Survival analysis showed that disease-free survival (DFS) of patients with low-level dendritic cell was longer than that of the high-level group in the TCGA cohort (Fig. 5e). Similar results were observed in macrophages (Supplementary Fig. 5a). However, low-level neutrophil was observed to be associated with better prognosis (Supplementary Fig. 5b). These results suggested that macrophages, dendritic cells, and neutrophils are more relevant to the development of PTC.

**Fig.5.**
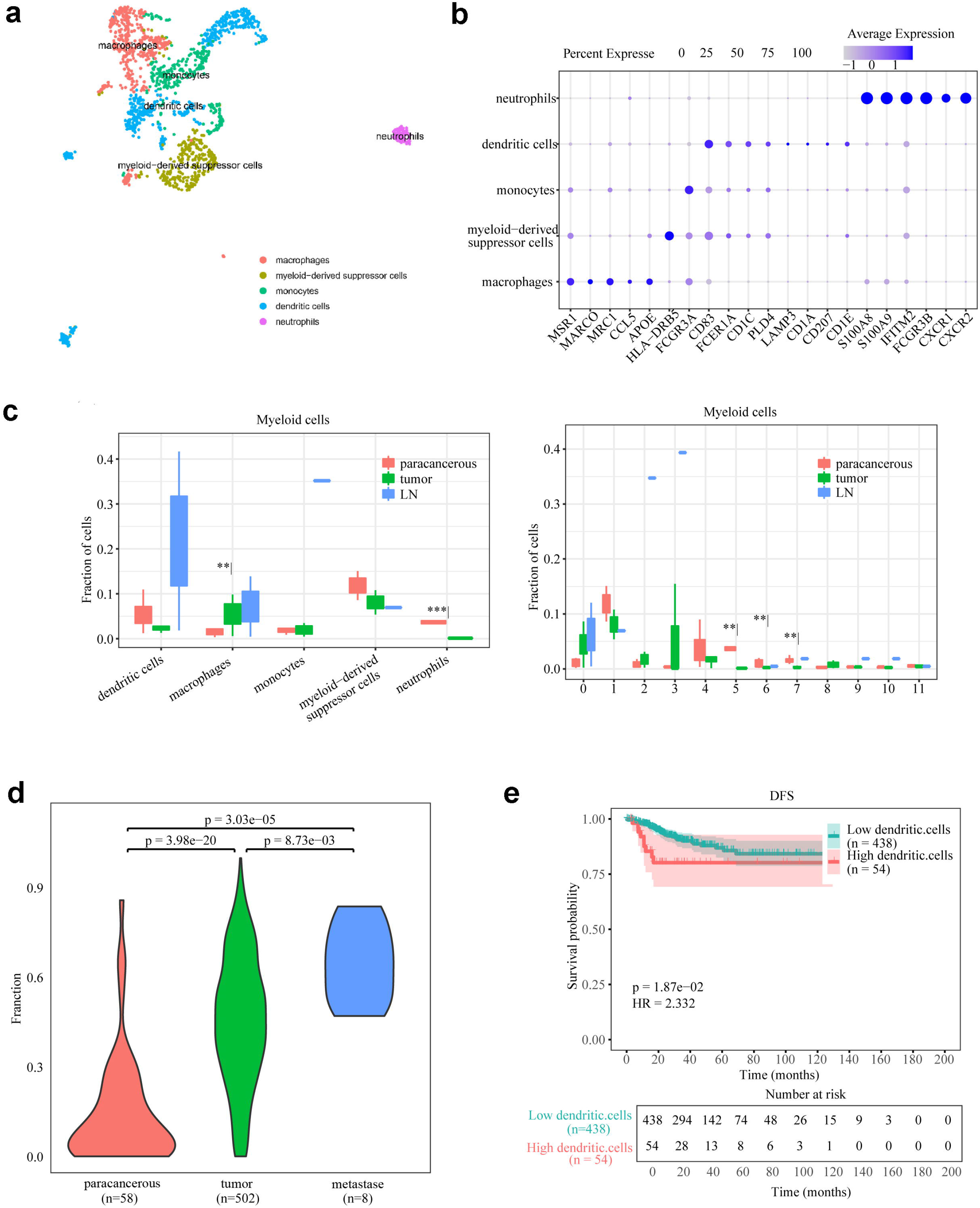
Heterogeneity of Myeloid cell populations. **a** UMAP plot of Myeloid cells color-coded by their associated cell types. **b** Dotplot of cell markers; sizes of dots represent abundance while color represents expression level. **c** Changes in frequency of multiple cell types and clusters in Myeloid cell populations. Asterisks on the left of the vertical line denote statistically significant differences between tumors and paracancerous tissues while asterisks on the right are used to show statistically significant differences between LNs and tumors. *p < 0.05, **p < 0.01, ***p < 0.001, two-tailed t-tests. **d** Violin of Macrophage abundance predicted from the TCGA THCA cohort by CIBERSORTx. **e** Kaplan-Meier curves of DFS based on the percentage of dendritic cells in the TCGA database.

We further characterized the functions of myeloid cell subtypes by comparing pathway activities. Compared with paracancerous tissues, the cancer hallmark-related pathways were relatively enriched in myeloid-derived suppressor cells from tumors, whereas they were generally down-regulated in neutrophils from tumors (Fig. 6a). These were consistent with the changes in cell proportions we observed earlier (Fig. 5c). Compared with tumor tissues, however, some cancer hallmark-related pathways (i.e. mTORC1 signaling and E2F targets) were down-regulated in myeloid-derived suppressor cells from metastatic LNs (Fig. 6b). The results indicated that myeloid-derived suppressor cells were involved in cancer cell proliferation, but not in tumor metastasis.

**Fig.6.**
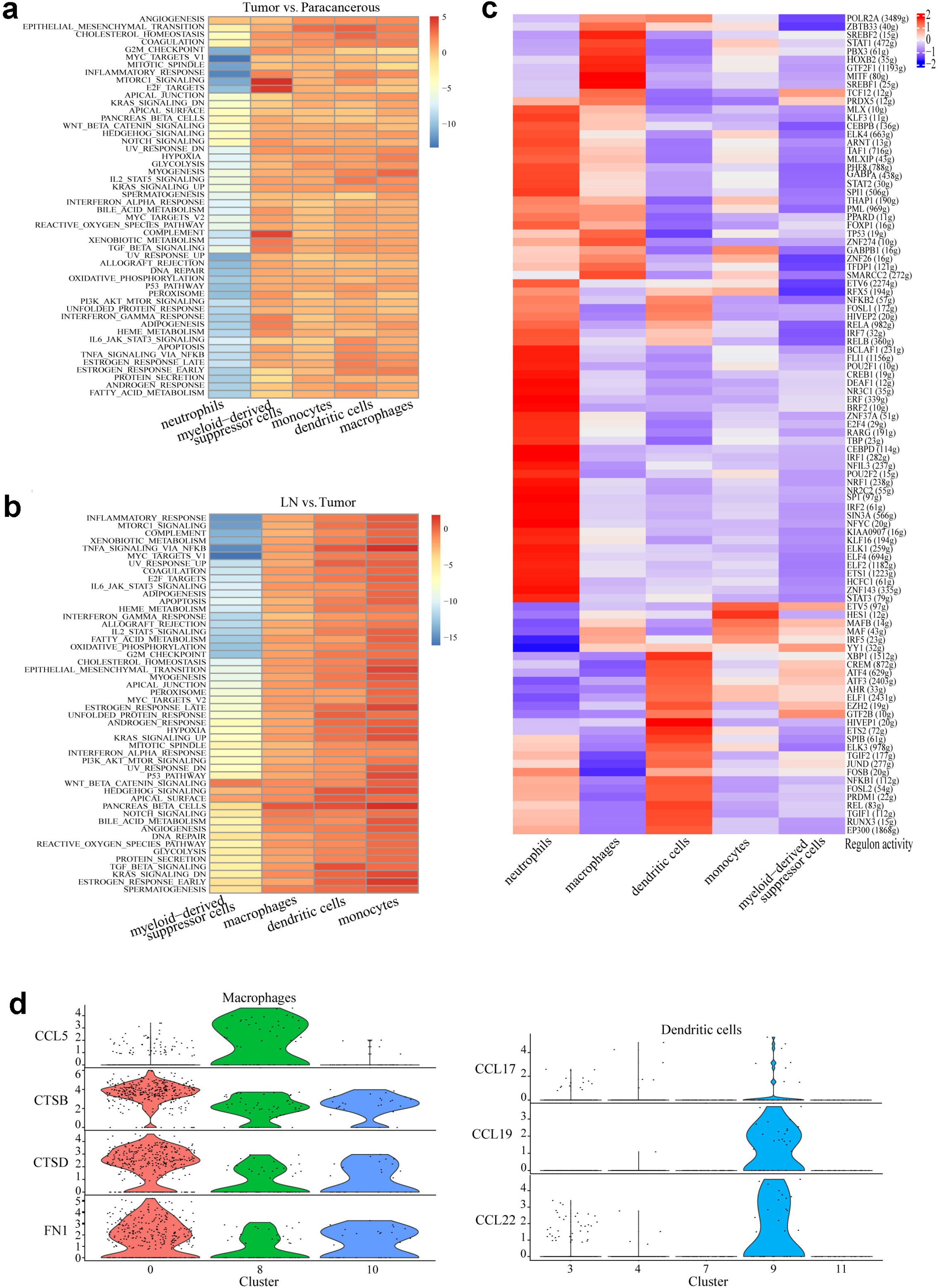
M2 macrophages were strongly enriched in the tumor tissues. **a-b** Heatmap shows the differences in pathway activities scored by GSVA between different sample origins. Shown are t values calculated by a linear model. **c** Heatmap of the AUC scores of TF motifs estimated by SCENIC. Shown are differentially activated motifs in subtypes of Myeloid cells, respectively. **d** Violin plot shows the expression level of related genes in Macrophage subclusters (left) and cytokines in dendritic cells (right).

In order to screen for key genes related to tumor occurrence and development in myeloid cells, we conducted SCENIC analysis and identified essential motifs in myeloid cells subtypes. Similar to the results of pathway analysis, the overall regulatory factors in neutrophils were also inconsistent with the other cell types (Fig. 6c). We recognized that the decreased activity of a lot of motifs (i.e. YY1 and IRF5) and activation of the STAT3, ZNF143, and HCFC1 motifs led to the reduction of neutrophils.

As shown in Fig. 6d, cells from myeloid cell-derived cluster 8 uniquely expressed the M1 macrophages marker CCL5, whereas M2 macrophages markers including CTSB/CTSD/FN1 were relatively highly expressed in cluster 0. The proportion of cluster 0 was high in tumors and LNs, although the difference has not reached a significant level (Fig. 5c). All of this evidence certified that cluster 0 represented an M2-like tumor-associated macrophages (TAM) cluster, the increase of which may be related to disease progression. Cluster 9 highly expressed the cytokines CCL17/CCL19/CCL22. These cytokines could bind to CCR4, a marker on the cell membrane of CD4^+^ Tregs, and showed strong chemotaxis to CD4^+^ Tregs ^37, 38^. It corroborated with our above results that the increasing trend of CD4^+^ Tregs with disease progression was coincident with that of myeloid cell-derived cluster 9 (Fig. 3f and 5c). The findings indicated that the dendritic cells (cluster 9) could recruit CD4^+^ Tregs into the tumor regions, thereby acting as an immunosuppressive effect.

## Discussion

At present, the study of molecular mechanisms underlying the occurrence and development of PTC mainly focused on the genetical alteration based on bulk sequencing data. We profiled integrated and heterologous transcriptional landscapes of PTC using single-cell sequencing methods, including cancer cells and TME of primary tumors and metastatic lesions. Through sequencing 28,205 cells, we found that the majority of cell clusters possessed strong heterogeneities. Tumor heterogeneity exists in various malignancies and remains not only between tumors but also within a single tumor ^39^, which is considered as a cause of chemoresistance of cancer. In-depth knowledge of tumor heterogeneity of PTC will make molecular testing more reliable and accurate prior to surgery (whether to undergo the surgery or close follow-up only), and benefit to stratify the recurrence risk and personalized precision treatment (thyroidectomy or total thyroidectomy or plus radioiodine treatment). On the other hand, TME on disease progression and metastasis has also been confirmed in numerous diseases^40, 41^. Investigations of TME-related cellular and molecular events will provide a theoretical rationale for drug discovery and development, especially for targeted therapy and immunotherapy.

In addition to previously described cell types, we discovered a novel cell subpopulation, named Protective EGR1^+^CD4^+^T cells. EGR1 is a transcription factor primarily mediate cellular functions (i.e. cell growth, cancer progress and apoptosis) via the RAS/RAF/MEK/ERK signaling pathway ^42^, which was a hallmark of PTC ^43^. It has also been previously reported that p53 could bind to EGR1 promoter and subsequently resulted in efficient apoptosis ^44^. EGR1 was usually considered a tumor suppressor in many human malignancies including PTC ^45, 46, 47, 48^. By comparing the proportion of cells from different tissues and pathway enrichment analyzing, we found that KRAS signaling pathway (part of RAS/RAF/MEK/ERK signaling pathway) was significantly down-regulated in Protective EGR1^+^CD4^+^ T cells. It indicated that Protective EGR1^+^CD4^+^T cells might inhibit the formation of the immunosuppressive microenvironment and tumor immune evasion via RAS/RAF/MEK/ERK signaling pathway, thereby restraining tumor growth. Therefore, EGR1 might represent a potential therapeutic target for PTC.

CD4^+^ Tregs, were overall considered to disrupt anti-tumor immunity and then help tumor cells to achieve immune evasion, leading to the tumor growth and metastasis ^49^. Our results also showed that CD4^+^ Tregs had the greatest positive correlation with the process of EMT, which matched those reported in other studies of PTC patients ^50^. Using pathway enrichment analysis, we found that relevant pathways associated with CD4^+^ Tregs and Protective EGR1+CD4+T cells were almost the opposite. The proportion of CD4^+^ Tregs among three samples increased in the disease progression order (paracancerous tissues < tumors < LNs) while the proportion of Protective EGR1^+^CD4^+^T cells decreased sequentially. This strongly suggested that CD4^+^ Tregs and Protective EGR1^+^CD4^+^T cells might act antagonistically to each other.

Immunotherapy is an emerging method for cancer treatment and promising results have been obtained in both hematologic and solid tumors ^51, 52^. T cell exhaustion is a critical mechanism of immune evasion. Blockade the PD1/PDL1 interaction to reverse T cell exhaustion is considered a milestone achievement in the field of immunotherapy ^53, 54, 55^. Our results showed that PDCD1 (PD-1), TIGIT, and CD96 could be suitable targets of immunotherapy in PTC patients without LN metastasis since they were upregulated in tumor tissues. However, the expression levels of TIGIT and CD96 were much higher than that of PDCD1 (PD-1) in LN-derived exhausted T cell types (Exhausted CD4^+^ T cells and Pre-exhausted CD8^+^ T cells). Therefore, TIGIT and CD96 might be better therapeutic targets for immunotherapy in PTC patients with LN metastasis than PDCD1 (PD-1). Jill et al. ^56^ investigated the CD4^+^ and CD8^+^ T cell exhaustion in PTC patients with LN metastasis using flow cytometry, and found that CD8+ T cell exhaustion was incomplete. This was most likely because TIGHT and CD96 were not included into analysis in their study, both of which were very important inhibitory immune checkpoints. However, their findings supported our conclusions to some extent that PDCD1 (PD-1) was not a suitable molecular target for the treatment of PTC with LN metastasis.

We initially applied a commonly used method inferCNV for the identification of malignant cells ^57^, which was shown to be unable to identify efficiently malignant tumor cells from epithelial cells in this study. Thus, we adapted an integrative Bayesian segmentation approach called CopyKAT and solved the conundrum successfully ^35^. As a result, more than half of tumor-derived epithelial cells were non-malignant cells, which might well explain the reason for the indolent clinical behavior of PTC. Additionally, there were also a very small number of malignant epithelial cells in paracancerous tissues, possibly due to the presence of the pre-tumor microenvironment in paracancerous tissues. There were several possible reasons. First, the pre-metastatic niche, a favorable microenvironment to tumor metastasis, formed in paracancerous tissues ^58^, and in turn lead to intra-thyroid metastasis. The concept of intra-thyroid metastasis of thyroid cancer has not been described yet. The reason is possibly caused by the small size of the thyroid gland itself and the narrow lumen of artery/vein nourishing the thyroid gland. As is well-known, liver cancer tends to present with intrahepatic metastatic disease, mainly due to the large diameter of the portal vein and hepatic artery. Second, multifocal PTC has already existed within the thyroid gland, whereas the extremely tiny lesion cannot be detected under microscopic examination. Based on the two points above, thyroidectomy is necessary to be performed for patients with PTC at least to ensure a total removal of potential lesions. Third, CopyKAT is not able to discriminate malignant cells from non-malignant cells with hundred percent accuracy.

In summary, we initially investigated the single-cell level heterogeneity of primary PTC tumors and metastatic lesions. A novel cell type, named Protective EGR1+CD4+T cells, was identified, which might inhibit the formation of the immunosuppressive microenvironment and tumor immune evasion. TIGIT and CD96 might be better therapeutic targets for immunotherapy in PTC patients with LN metastasis than PDCD1 (PD-1). These findings would contribute to the further understanding of molecular mechanisms resulting in occurrence and development of PTC, and provide a theoretical rationale for targeted therapy and immunotherapy.

## Methods

### Sample collection and clinical information

Seven patients diagnosed with PTC were obtained from Shanghai Tenth People’s Hospital. A total of 15 tissue samples (7 tumor tissues, 6 paracancerous tissues, and 2 metastatic LNs) were obtained from 7 PTC patients. Written informed consent was obtained from all patients in the present study. Histological diagnosis of all the samples was confirmed by the pathology department.

### Tissue digestion and single-cell suspension preparation

Cells of each sample were firstly stained with two fluorescent dye, Calcein AM (Thermo Fisher Scientific Cat. No. C1430) and Draq7 (Cat. No. 564904), for precisely determination of cell concentration and viability via BD Rhapsody™ Scanner before single-cell multiplexing labeling. The cells of each sample were sequentially labeled with BD Human Single-Cell Multiplexing Kit (Cat. No. 633781) which utilizing an innovative antibody-oligo technology ^22^ mainly to provide higher sample throughput and eliminate batch effect for single-cell library preparation and sequencing. A set of 12 antibodies in this kit recognize the same universally expressed cell-surface antigen of human cells. Each antibody is conjugated with a Sample Tag, a unique 45-nucleotide barcode sequence. Briefly, cells from each sample were labeled by antibodies with different sample tags respectively and washed twice with BD Pharmingen™ Stain Buffer (FBS) (Cat. No. 554656) before pooling all samples together. BD Rhapsody Express system^23^based on Fan et al. was utilized for single-cell transcriptome capturation. Pooled samples were then loaded in one BD Rhapsody™ Cartridge that was primed and treated strictly following the manufacturer’s protocol (BD Biosciences). Cell Capture Beads (BD Biosciences) were then loaded excessively onto the cartridge to ensure that nearly every micro-well contains one bead, and excess beads were washed away from the cartridge. Viable cells with beads captured in wells were detected in BD Rhapsody™. Then cells were lysed, Cell Capture Beads were retrieved and washed before performing reverse transcription and treatment with exonuclease I.

### Library construction and sequencing

Transcriptome and SampleTag information of single-cells was obtained through BD Rhapsody System. Microbeads-captured single cell transcriptome and SampleTag sequences were generated into the cDNA library and SampleTag library separately containing cell labels and unique molecular identifiers (UMI) information. Briefly, double-strand cDNA was firstly generated from microbeads-captured single-cell transcriptome through several steps including reverse transcription, second-strand synthesis, end preparation, adapter ligation, and whole transcriptome amplification. Then final cDNA library was generated from double-strand full-length cDNA by random priming amplification with BD Rhapsody cDNA Kit (BD Biosciences, Catalog No.: 633773) and BD Rhapsody Targeted mRNA & AbSeq Amplification Kit (BD Biosciences, Catalog No.: 633774). On the other hand, the SampleTag library was generated from microbeads-captured single-cell SampleTag sequences through several steps including reverse transcription, nest PCR, and final index PCR. Libraries were sequenced on the NovaSeq platform (Illumina).

### Raw data preprocessing and quality control

Raw sequencing reads of the cDNA library and SampleTag library were processed through the BD Rhapsody Whole Transcriptome Assay Analysis Pipeline (Early access), which included filtering by reads quality, annotating reads, annotating molecules, determining putative cells, and generating single-cell expression matrix. Briefly, read pairs with low sequencing quality were firstly removed. The quality-filtered R1 reads were analyzed to identify the sequence of cell labels, UMI sequence, and poly-dT tail sequence, meanwhile the quality-filtered R2 reads were mapped to Genome Reference Consortium Human Build 38 (GRCh38) using STAR (version 2.5.2b) in the reads annotation step. Further adjustment was performed with the recursive substitution error correction (RSEC) and distribution-based error correction (DBEC) algorithms to remove artifact molecules due to amplification bias in the molecules annotation step. Putative cells were distinguished from background noise with second derivative analysis in the putative cell determination step. Finally, putative cells information was combined with molecules adjusted by the recursive substitution error correction and distribution-based error correction algorithms to generate a single-cell expression matrix. The pipeline also determined the sample origin of every single cells via the sample determination algorithm according to the sequencing reads of the SampleTag library. The algorithm classified all putative single cell into three categories: Multiplet, two or more SampleTag exceed their minimum thresholds, indicating more than one actual cell in one micro-well. Undetermined, not enough SampleTag reads for the sample origin. SampleTag01-12, only one SampleTag exceed their minimum thresholds. . Among all the output files, Matrix for UMI counts per cell corrected by DBEC and SampleTag annotation result were used for downstream clustering analysis.

### Dimension reduction and clustering

The R package Seurat ^23^ was utilized for subsequent analysis. Raw gene expression matrices from the cartridge were read into R (version 3.6.0) and converted to Seurat objects. Cells label as “Undetermined” and “Multiple” were excluded in the following analysis. The gene expression matrix was then normalized to the total cellular UMI count. Top 2000 features were selected as highly variable genes for further clustering analysis. In order to reduce dimensionality, PCA was performed based on the highly variable genes after scaling the data with respect to UMI counts. On top of that, the first 9 principal components were chosen for downstream clustering based on the heatmap of principle components, and the elbow plot of principle components to further reduce dimensionality using the UMAP algorithm. The transcriptional markers of each cluster were calculated using the FindAllMarkers function with theMAST package to run the DE testing under the following criteria: log2 fold change > 0.1; p < 0.05; min. percentage > 0.25. Top 500 markers of each cluster were then selected to perform a heatmap plot.

### Cell annotation and cell type identification

Cell populations were matched to cell types based on the expression of known marker genes and previously identified expression signatures ^23, 24, 25, 26, 27, 28^.

### Comparison of cell clusters and cell type proportion

The change in the fraction of the different cell types was separately computed for each sample across all clusters, as the fraction of cell in each cluster, out of the total number of cells ^29^. To assess a statistically significant difference in a fraction of a specific cell type, we performed paired t-test.

### Significantly dysregulated genes identification

We identified differentially expressed genes (DEGs) based on analysis of the MAST package using the R. The threshold of DEGs was set as: 1)P-value of F test < 0.05; 2) |FC| ≥1.5.

### External data validation with TCGA datasets

CIBERSORTx was used to detect the abundance of cell types identified in the present work in bulk RNA-seq data ^30^. After calculated the relative abundance of each cell type, we divided the patients into two groups: high 50% and low 50%. Subsequently, Kaplan-Meier analysis was performed using ggsurvplot function of the R package ‘survminer’. Whether the occurrence of cell types in the different subgroups showed significant differences was evaluated by Wilcoxon test.

### Functional annotation and enrichment

GO enrichment and KEGG enrichment of DEGs were performed using Fisher exact test with Benjamini-Hochberg multiple testing adjustment. The results were visualized using R package. Gene set variation analysis (GSVA) ^31^ was performed using 50 hallmark-related gene sets, as described in the GSVA package.

### Identification of hub regulons with SENIC

In order to investigate the gene regulatory network of different sample groups, we utilized SCENIC ^32^ using the R.

### Cell-cell communication analysis based on ligand-receptor pairs

Cell-cell communication at the molecular level was analyzed with CellPhoneDB ^33^.

### Cell trajectory analysis with Monocle

In order to reveal the development of malignant cells, we employed Monocle 2, an R package designed for single cell trajectories ^34^. After obtaining the differentially expressed genes through differential Gene Test function, the trajectories were visualized as 2D tSNE plots.

### Drug-target analysis with TCMID database

After obtaining the differentially expressed genes, we obtain the targeted relationship between drugs and genes from the TCMID database (http://www.megabionet.org/tcmid).

### CNV Estimation

CopyKAT ^35^ (Copynumber Karyotyping of Tumors), a R package was performed to separate tumor cells from normal cells using high-throughput sc-RNAseq data. Cells with extensive genome-wide copy number aberrations (aneuploidy) are considered as tumor cells, whereas stromal normal cells and immune cells often have 2N diploid or near-diploid copy number profiles.

### Statistical analysis

Box plots were drawn with the R base package. Hence, the boxes span from the 25th to the 75th percentiles. Violin plots were generated by the ggplot2 R package. Unpaired two-tailed t-tests was used to compare the difference between two groups. While One-way analysis of variance (ANOVA) was used for multiple group comparisons.

## Data Availability

The raw data can be obtained with permission from the corresponding author.

## Data availability

The cancer genome atlas (TCGA) thyroid cancer (THCA) cohort were downloaded from UCSC XENA (http://xena.ucsc.edu/).

## Acknowledgments

This work was supported by Shanghai Municipal Commission of Health and Family Planning Commission (2019SY062).

## Authors’ contributions

BH J, ZS W, ZQ Y, and Y R conceived and designed the study. WL J, CG T, and ZS W contributed to carry out the experiments. BH J, CG H, and C Y contributed to data analysis. ZS W, BH J, and ZQ Y wrote the manuscript. YR, WL J, and ZQ Y revised the paper. All authors read and approved the final manuscript.

## Supplementary information

**Supplementary Fig. 1 Identification of major cell types in PTC**. UMAP plots for expression of the marker genes from **Fig 1c** and of additional marker genes for major cell types. For B cells: CD19, MS4A1(CD20), CD38, CD79A, and CD79B; for CD4^+^ T cells: CD4 and CD3D; for CD8^+^T cells: CD8A and CD3D; for Endothelial cells: CD34 and CD31(PECAM); for Epithelial cells: EPCAM and KRT18; for Fibroblasts: COL1A1; for Myeloid cells: CD14, CD86, ITGAX, CD80, CD83, and ITGAM; for Naive T cells: CCR7 and CD3D; for NKT cells: NKG7 and CD3D; for Plasma cells: CD79A and SDC1.

**Supplementary Fig. 2 Overview of the 28**,**205 single cells from tumors, paracancerous tissues, and metastatic LNs. a** The fraction of multiple cell types originating from 7 tumor tissues, 6 paracancerous tissues, and 2 metastatic LNs. **b** The fraction of multiple cell types originating from 15 samples. **c** The number of multiple cell types. **d** Box plots of the number of transcripts.

**Supplementary Fig. 3 Potential new cell markers in T cells**.

**Supplementary Fig. 4 DEGs related to CD4**^**+**^**Tregs and transitional EGR1**^**+**^**CD4**^**+**^**T cells. a** The expression of the maker genes in the TCGA THCA cohort. **b** Differences in 50 hallmark pathway activities scored with GSVA software. Shown are t values calculated by a linear model. **c** DEGs related to Protective EGR1^+^CD4^+^ T cells in tumors and LNs, respectively. **d** DEGs related to CD4^+^Tregs in tumors and LNs, respectively. Upregulated genes and downregulated genes were colored in red and blue, respectively. FC≥2.

**Supplementary Fig. 5 External verification based on the TCGA data. a** High level of Macrophage abundance predicted poor prognosis in the TCGA THCA cohort, and neutrophil was the opposite. **b** Kaplan-Meier survival curve for IFITM2.

