## Supplementary figures and images for "Single-cell RNA sequencing reveals a novel cell type and immunotherapeutic targets in papillary thyroid cancer"

### Supplementary Fig. 1

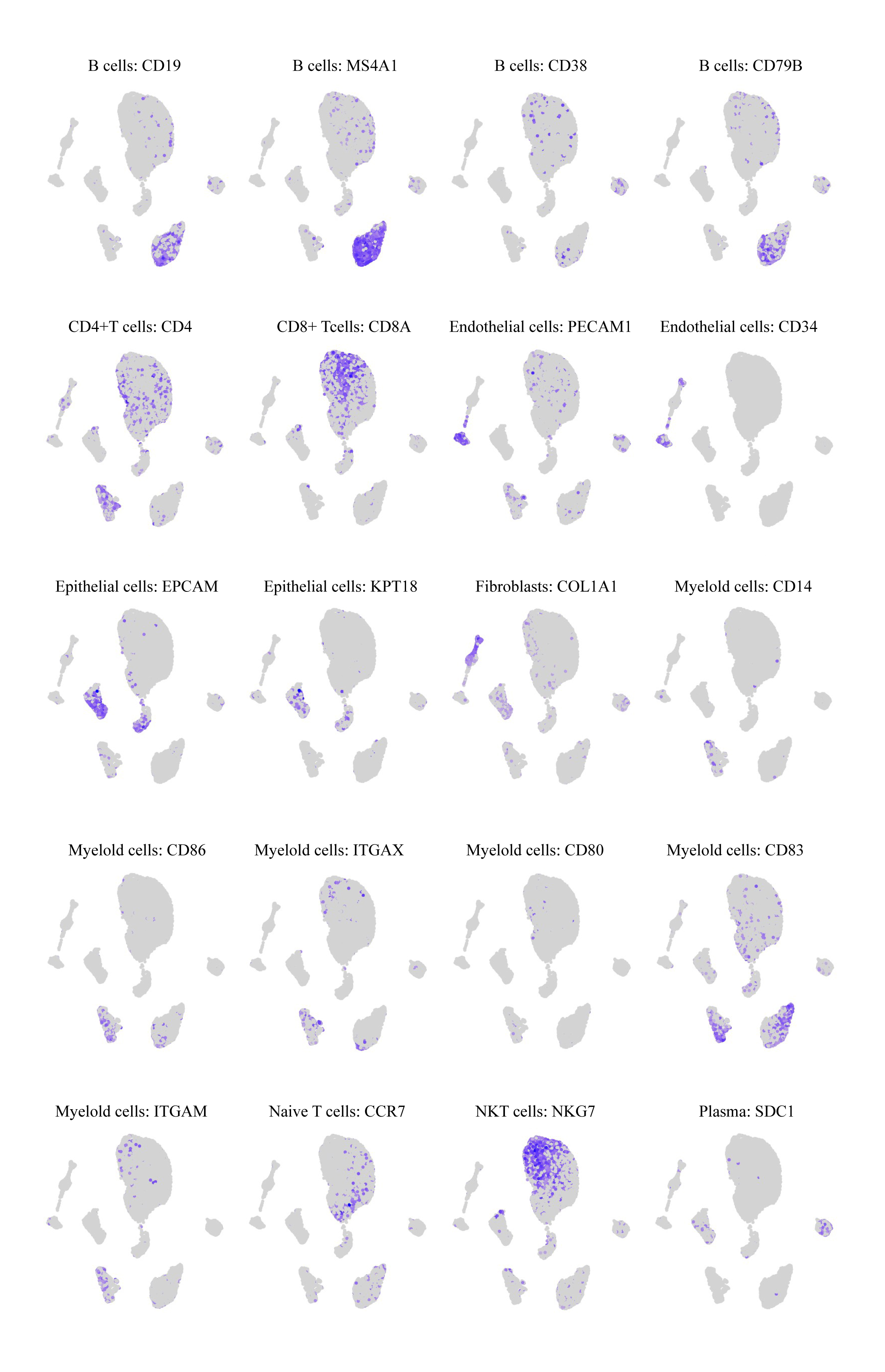

### Supplementary Fig. 2

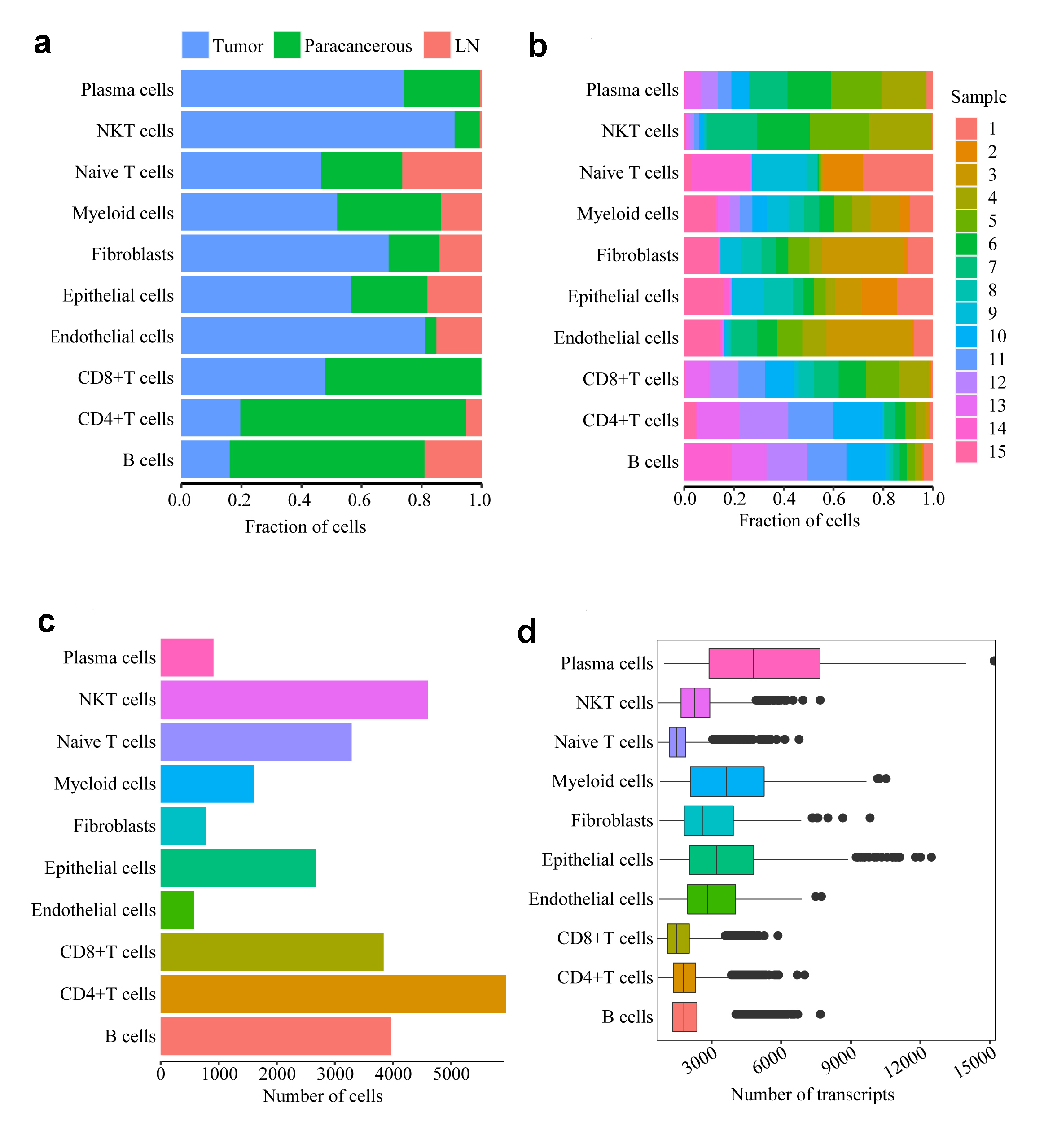

### Supplementary Fig. 3

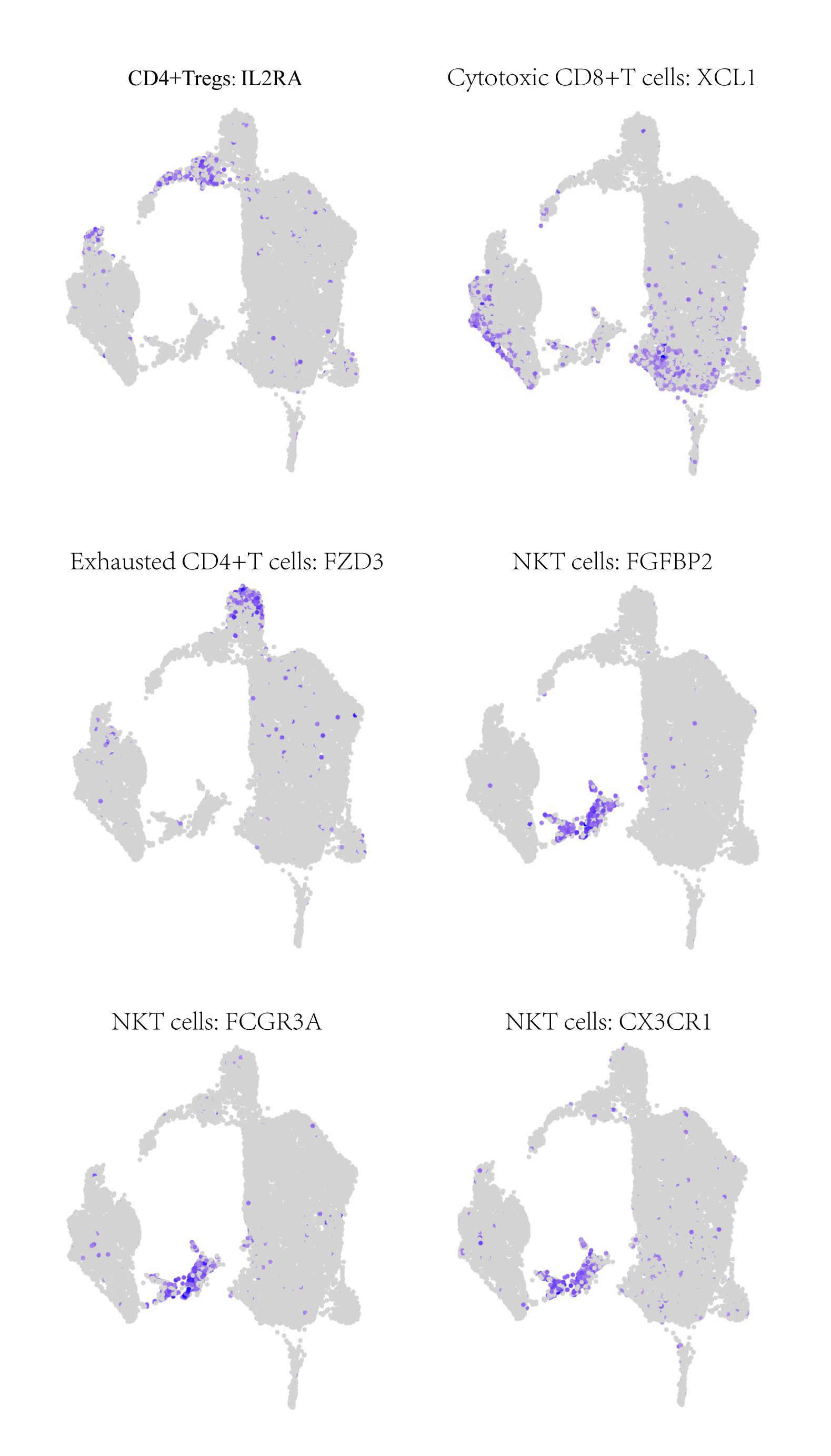

### Supplementary Fig. 4

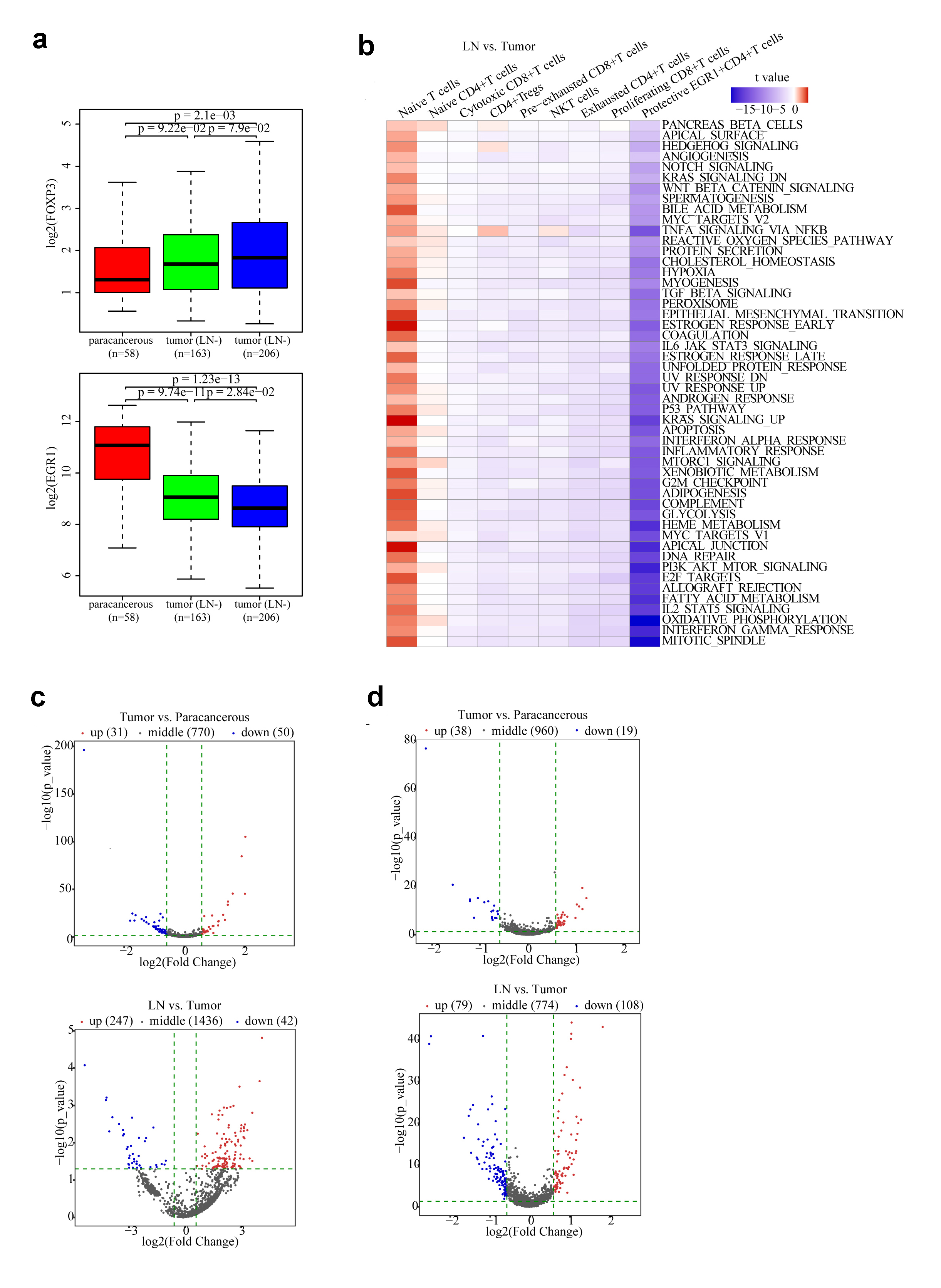

### Supplementary Fig. 5

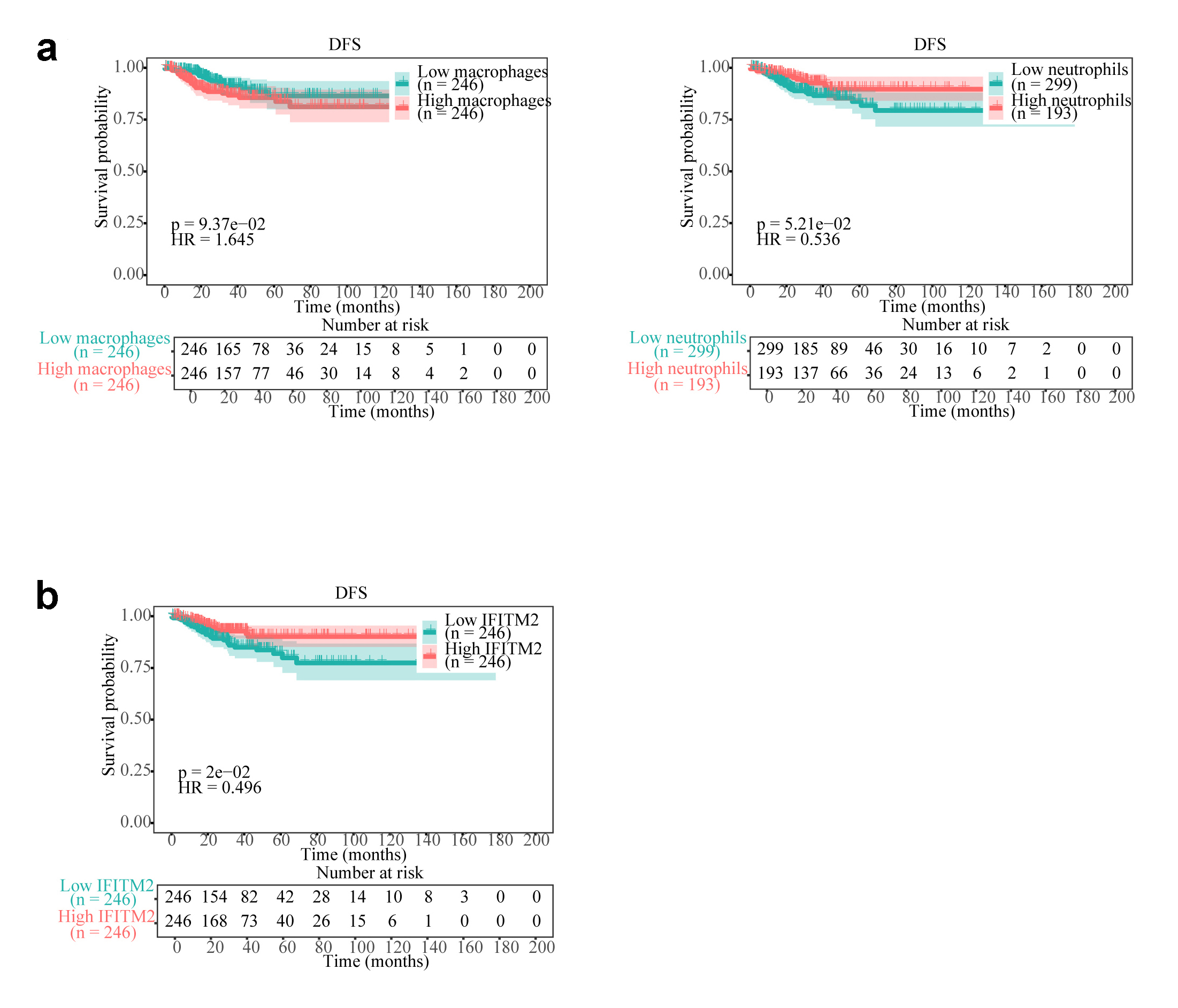
